# An Evaluation of The Regional Distribution of Pediatric Surgery Workforce and Surgical Load in Brazil

**DOI:** 10.1101/2022.10.19.22281261

**Authors:** Joaquim Bustorff-Silva, Márcio Lopes Miranda, Amanda Rosendo, António Gonçalves de Oliveira Filho

**Affiliations:** Division of Pediatric Surgery, Department of Surgery, State University of Campinas Medical School, Rua Tessália Vieira de Camargo, 126. Cidade Universitária Zeferino Vaz. CEP 13083-887 – Campinas, SP, Brasil

**Keywords:** Pediatric Surgery, Workforce, Workload, Brazil

## Abstract

**Aim:** The purpose of this study is to present data on the regional distribution of the pediatric surgical workforce and the expected local demand of pediatric operations in Brazil.

**Methods:** We collected data on the pediatric surgical workforce, surgical workload, GDP/capita and mortality for gastrointestinal tract malformations (MGITM) across the different regions of Brazil for 2019.

**Findings:** Data from CFM reported the existence of 1515 pediatric surgery registries in Brazil, corresponding to 1414 individual pediatric surgeons (some pediatric surgeons are registered in more than one State), or 2.4 pediatric surgeons per 100.000 children< 14 years. There were 828 male and 586 female with a M/F ratio of 1.14:1. Mean age was 51.5±12.8 years and mean time from graduation was 3,4±5,7years. Regional distribution showed a higher concentration of pediatric surgeons on the wealthier Central-West, South and Southeast regions. Individual workload ranged from 88 to 245 operations/year (average 146 operations/year) depending on the region. Only 9 (6.1%) of these were high complexity (including neonatal) operations. MGITM tended to be higher on the poorer North and Northeast regions of Brazil

**Conclusions:** We found wide disparities in the surgical workforce and workload across Brazil, related to socioeconomic status. Areas of increased surgical workforce were associated with lower MGITM. The average number of complex operations performed yearly by the individual pediatric surgeon was considerably low. Strategic investment and definition of health policies may be needed to improve the quality of care in the different regions of Brazil.

**Level of Evidence:** Retrospective review. Level III

## Introduction

In recent years there has been an increase in publications of general data on the demand and workforce of Pediatric Surgery in several countries^1-7^.

In Brazil, despite some recent publications^8-11^, information regarding the exact size of the pediatric surgery workforce, as well as the regional demands for pediatric operations is scarce and in most instances ambiguous, depending on the source of the information. This lack of reliable information makes it difficult to plan the need of these professionals across the different regions of Brazil and also to estimate the need for training new pediatric surgeons every year.

The purpose of this study is to present data on the regional distribution of the pediatric surgical workforce trying to correlate it with the expected local demand of pediatric operations and to present some potential alternatives.

## Methods

This study is an ecological, cross-sectional descriptive analysis using data from the Brazil public health system (Sistema Unico de Saude - SUS) and the Brazilian Institute of Geography and Statistics (IBGE).

Data from the IBGE from 2019, shows that Brazil had 213 million inhabitants and a Gross Domestic Product per capita (GDP/capita) of R$35,156.16 (Brazilian reais). It is composed by 26 States and a Federal District which are grouped in 5 major regions (North, Northeast, Southeast, South and Central-West). From the economic point of view, the North and Northern Regions are poorer, displaying an average GDP of approximately R$20,000.00 whereas the remaining regions display and average GDP of R$43,000.00.

To evaluate the pediatric surgery workforce and workload we consulted the databases available at IBGE (http://tabnet.datasus.gov.br/cgi/deftohtm.exe?ibge/cnv/popbr.def) taking as reference the year 2019, as well as the data from the Federal Medical Council (CFM) contained in the document MEDICAL DEMOGRAPHY IN BRAZIL 2020^12^. Pediatric surgeon workforce density (PSWD) was defined as the number of Pediatric Surgeons for each 100.000 children under 14 years of age. For the purpose of calculating the PSWD in this study we used data from this last publication because it derives data from CFM which is the official organ where doctors have to be registered to practice their specialty.

To estimate the surgical demand of the different states and regions of Brazil we used a pool of index surgical procedures modified from previously published data and classified it in two levels of complexity. Low complexity operations included hernia correction, orchidopexy, appendicitis, vascular access, correction of hypospadias, management of congenital megacolon, pyloromyotomy for pyloric stenosis and correction of pediatric gastroesophageal reflux. Complex operations included neonatal surgery (correction of intestinal atresia, anorectal malformation, esophageal atresia, abdominal wall defects, diaphragmatic hernia, neonatal enterocolitis), correction of biliary atresia and choledochal cysts and liver transplants. The expected number of surgeries was estimated based on the published incidence of each of the corresponding pathologies^9^. Surgeries performed because of trauma, oncologic surgeries, transplants or re-operations were not included in the demand estimate.

In order to have a rough evaluation of the outcomes of neonatal surgery in the different regions of Brazil we evaluated the mortality associated with a grouped diagnosis of Malformations of the Digestive System that included International Classification of Diseases – 10 (ICD10) codes from Q38 to Q45.

Data is presented in tables and graphs. Due to the descriptive nature of this study no attempt was made to do any statistical calculations with the data.

## Results

Data from CFM ^12^ reported the existence of 1515 pediatric surgery registries in Brazil, corresponding to 1414 individual pediatric surgeons (some pediatric surgeons are registered in more than one State), or 2.4 pediatric surgeons per 100.000 habitants younger than 14 years. Of these, 828 were male and 586 were female with a M/F ratio of 1.14:1. Mean age is 51.5±12.8 years and mean time from graduation is 3,4±5,7years.

Data from table 1 and figure 1 shows that the distribution of PS in Basil is not uniform. PSWD was higher in the Southeastern, South and Central-West regions of Brazil which tend to be the ones with the highest GDP/capita. North and Northeast regions have roughly half the concentration of Pediatric Surgeons than the other regions.

**Table 1.** Distribution of the demographic and Pediatric Surgery workforce and workload across the different regions of Brazil. (PS=Pediatric Surgeons)

| Demographic Data |  |  |  |  |  |  |
| --- | --- | --- | --- | --- | --- | --- |
| Region | North | Northeast | Southeast | South | Central-West | Brazil |
| Number of inhabitants (IBGE) | 18.906.962 | 57.667.842 | 89.632.957 | 30.402.587 | 16.707.336 | 213.317.684 |
| Number of live births | 301.635 | 770.688 | 2.093.142 | 796.536 | 230.474 | 4.192.475 |
| Childhood mortality (num/1000 births) | 16,6 | 15,3 | 11,7 | 10,1 | 13,0 | 13,3 |
| Number of children < 1 year old | 296.628 | 758.896 | 2.068.652 | 788.491 | 227.478 | 4.136.715 |
| Number of youngsters > 14 years (IBGE) | 3.483.144 | 15.397.314 | 23.932.000 | 8.117.491 | 4.460.859 | 56.955.822 |
| GDP per capita | 22.536,83 | 18.359,74 | 44.340,21 | 42.443,83 | 44.879,37 | 35.156,16 |
| Pediatric Surgery Workforce |  |  |  |  |  |  |
| Active PS | 65 | 256 | 796 | 259 | 139 | 1515 |
| Active PS/100000 youngsters | 1,9 | 1,7 | 3,3 | 3,2 | 3,1 | 2,7 |
| Number of youngsters/PS | 536 | 601 | 301 | 313 | 321 | 376 |
| Number of children > 1 year / PS | 4564 | 2964 | 2599 | 3044 | 1637 | 2731 |
| Expected number of operations / PS |  |  |  |  |  |  |
| Low complexity | 230,0 | 149,4 | 131,0 | 153,4 | 82,5 | 137,6 |
| High complexity | 15,1 | 9,8 | 8,6 | 10,0 | 5,4 | 9,0 |
| High Complexity except NEC | 5,9 | 3,9 | 3,4 | 4,0 | 2,1 | 3,5 |
| Total Yearly surgical load / PS | 245,5 | 159,5 | 139,8 | 163,8 | 88,0 | 146,9 |
| Weekly surgical load / PS | 5,1 | 3,3 | 2,9 | 3,4 | 1,8 | 3,1 |
| Mortality secondary to Gastrointestinal tract malformations |  |  |  |  |  |  |
| Number of admissions 2019 | 514 | 1317 | 2620 | 898 | 497 | 5846 |
| Number of deaths 2019 | 16 | 54 | 54 | 19 | 8 | 152 |
| Mortality rate (%) | 3,11 | 4,10 | 2,06 | 2,12 | 1,61 | 2,60 |

**Fig. 1.**
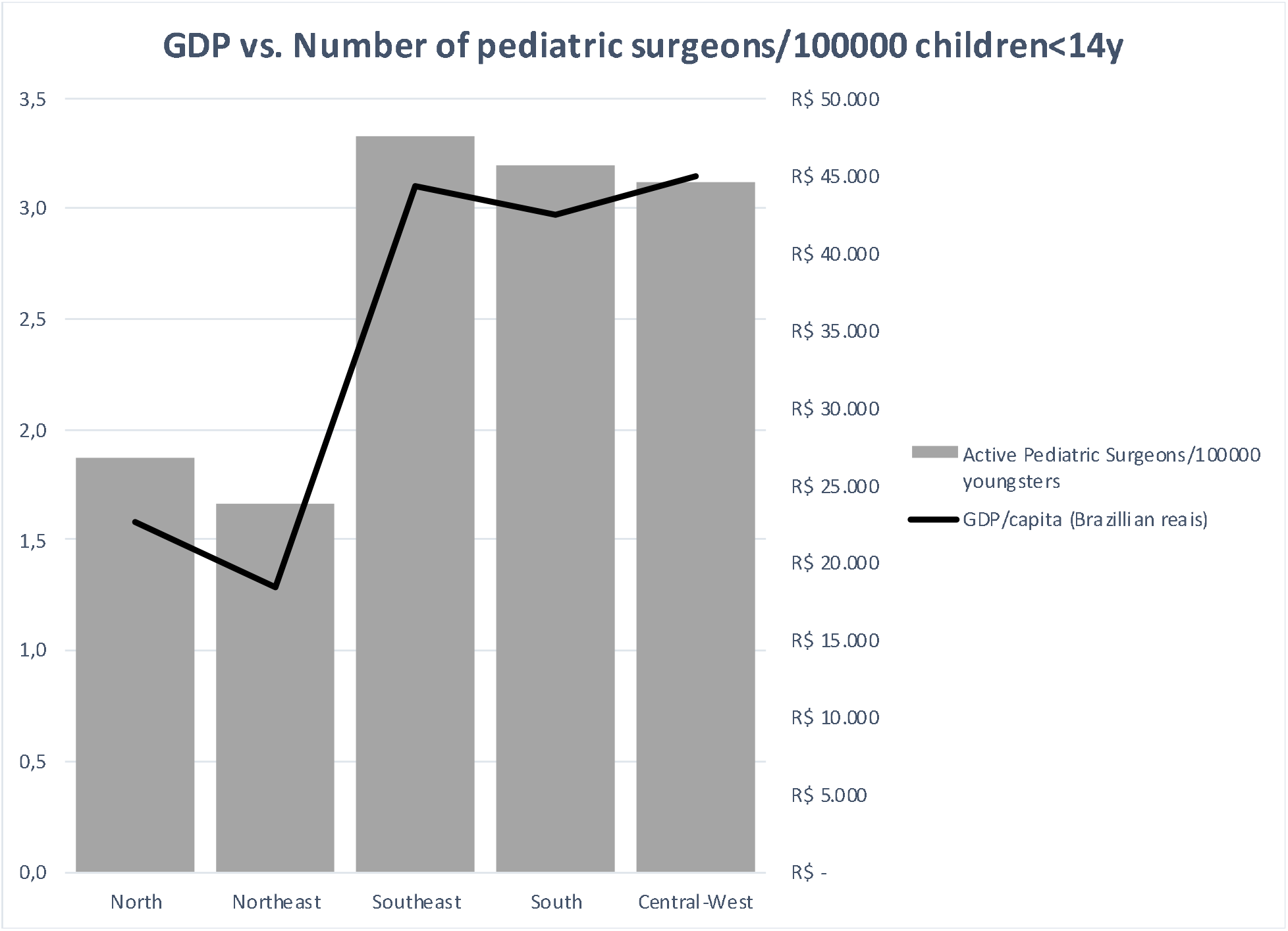
Graphic display of the distribution of the pediatric surgery workforce across the different regions of Brazil.

Accordingly, as the number of PS is lower in these two regions, the workload is greater than in the Central and Southern regions (figure 2). Based on the demographic data, a Brazilian pediatric surgeon is expected to perform an average of 147 pediatric operations a year or 3.1 a week. Of these operations only 9 (6.1%) fall into the high complexity operations. As can be seen in figure 3, for most neonatal operations the expected number is less than one/pediatric surgeon/year

**Fig. 2.**
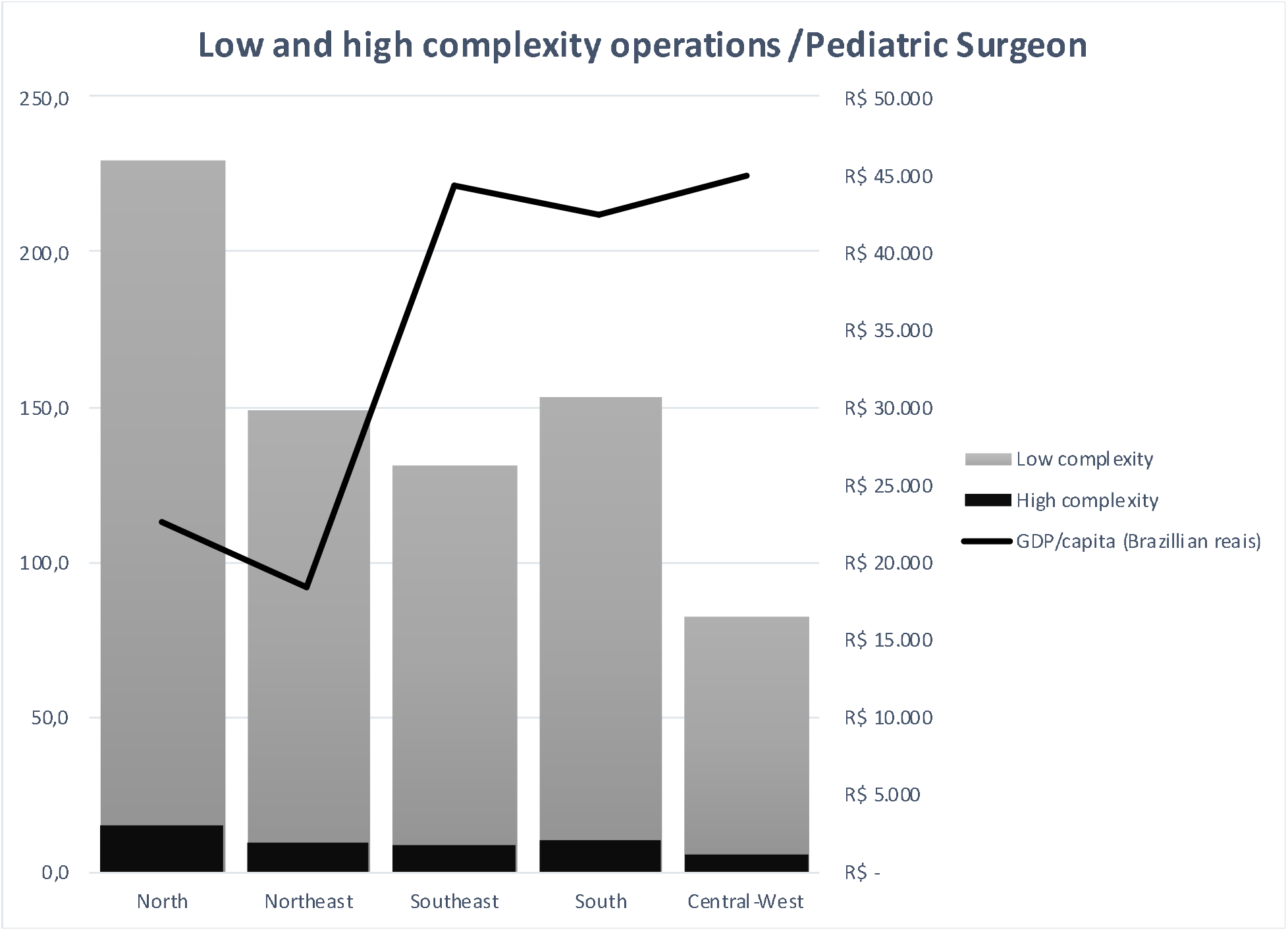
Graphic representation of the distribution of surgical workload of pediatric surgeons across the different regions of Brazil

**Fig. 3.**
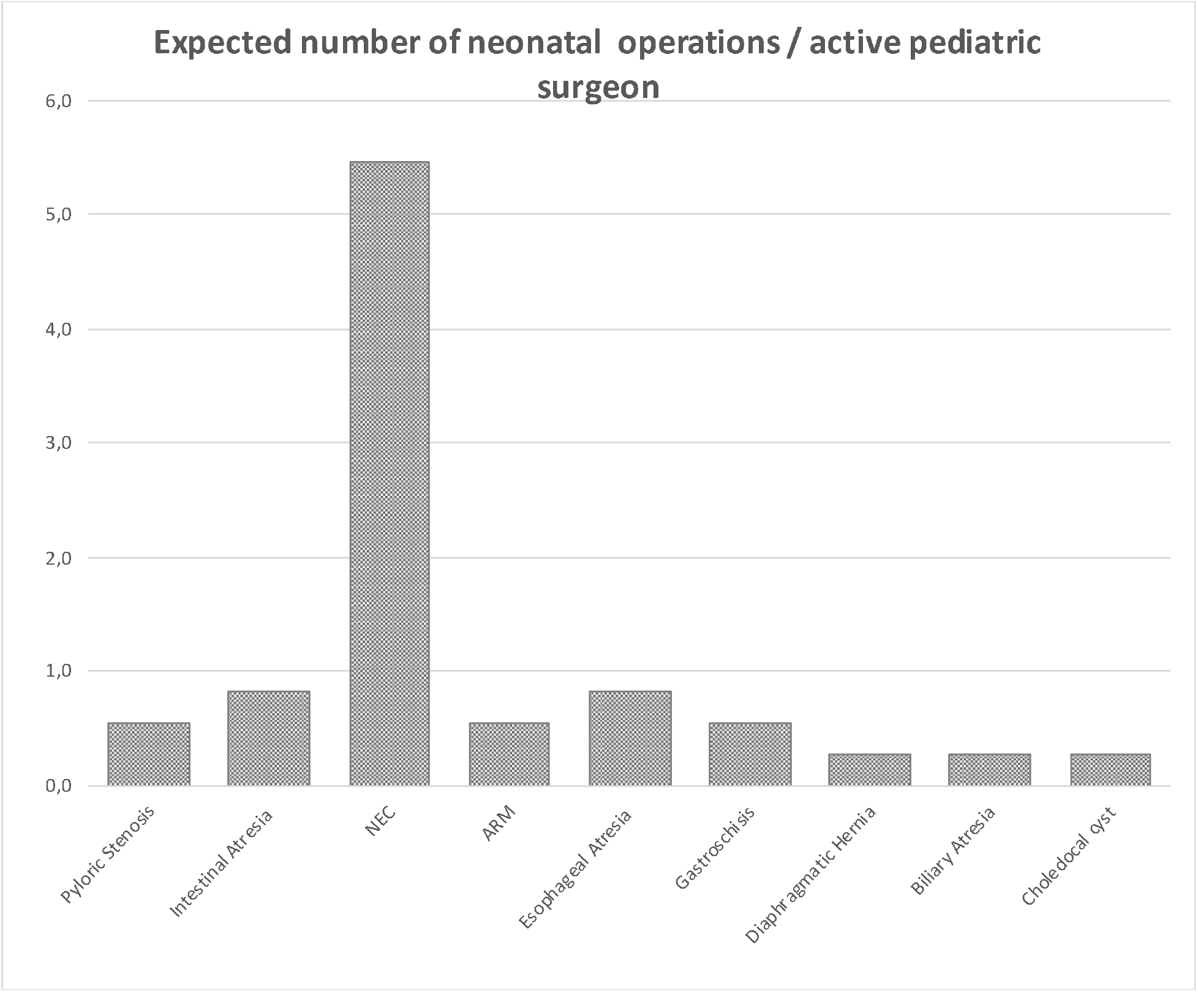
Graphic representation of the expected number of complex operations per pediatric surgeon per year, in Brazil

Figure 4 shows that the mortality rate of neonatal surgery in the Southern States (Southeast, South and Central-West) is roughly half of that observed in the North States (North and Northeast).

**Fig. 4.**
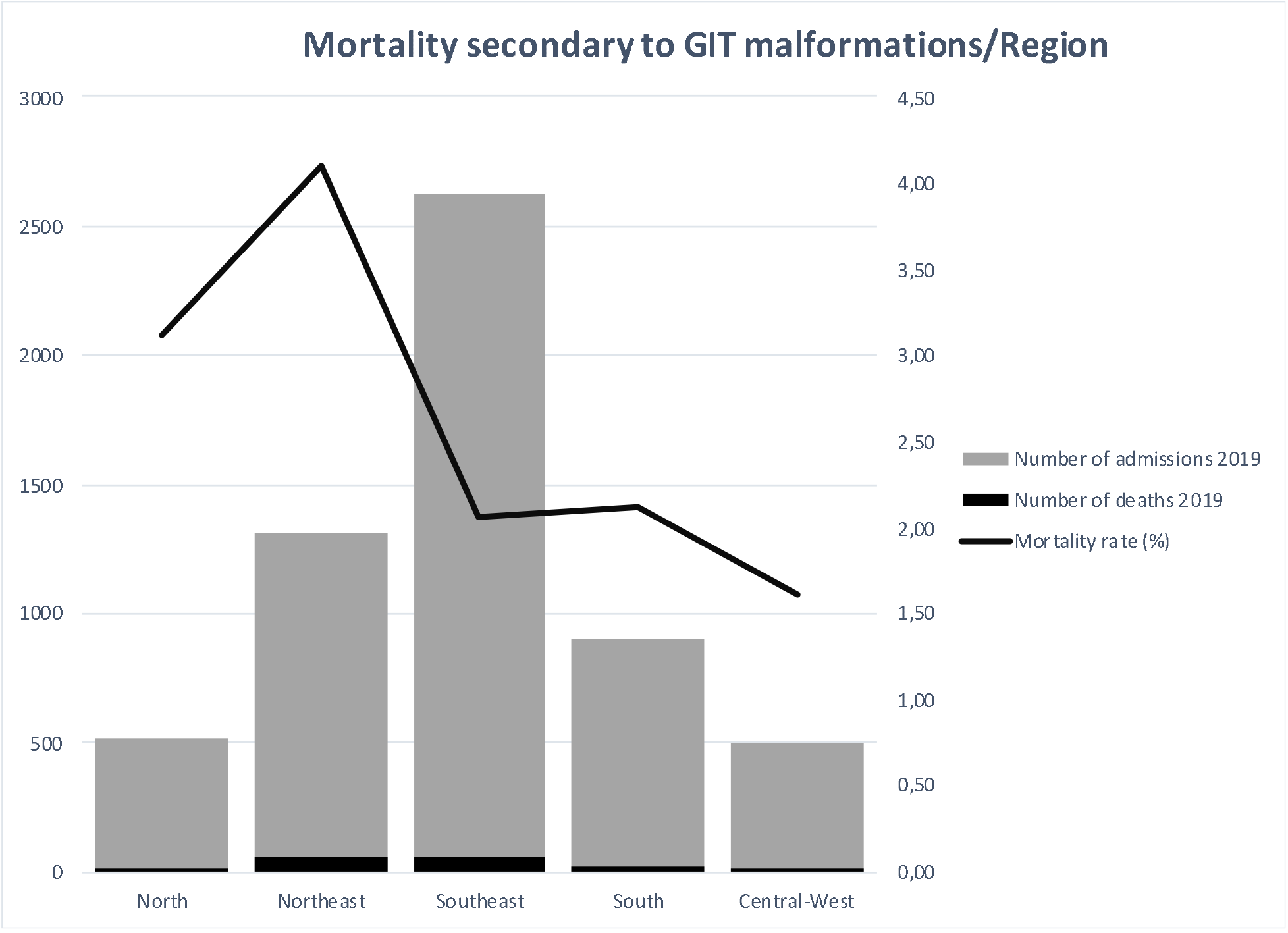
Graphic representation of the mortality secondary to Malformations of the Gastrointestinal Tract across the different regions of Brazil, in 2019.

## Discussion

Data from the present study shows that, in Brazil, Pediatric Surgery is a small surgical specialty when compared to other surgical specialties. The number of PS in Brazil represents about 0,31% of the total number of doctors in the country.

As stated previously, depending on the source of information, data may be confusing. In the year of 2019, while the CFM counted 1515 PS, the IBGE reported the existence of 674 Pediatric Surgeons in Brazil. This data disparity has been reported before but the cause is still unclear^9^.

Additionally, in 2019, the Brazilian Association of Pediatric Surgeons (CIPE) had only 454 registered members, meaning that about half of the surgeons who are operating on children in Brazil are not board certified by the CIPE. It is important to state that, theoretically, by Brazilian rules, one needs only a Federal Council of Medicine diploma to practice any medical or surgical specialty. However, although not mandatory, it is strongly believed that certification by a Medical Association (like CIPE) would assign a quality stamp to the professional activity of every pediatric surgeon.

Based on the CFM data, the Brazilian PSWD is slightly lower than the one observed in developed countries (2.65 vs. 3.28/100.000 children<14 years). Data from a European Census on Pediatric Surgery shows that in the European Union there are 3.9 PS/100.000 children and approximately 1662 neonates for each PS^13^. Middle and low income countries have respectively 9.4 and 5.1 PS/100.000 children^4^. Recent data suggests that there is a correlation between PSWD<0.37 with decreased odds of survival for specific pediatric conditions^1^. Accordingly, our findings reveal that mortality rate for congenital gastrointestinal tract malformations is higher in the lower PSWD regions.

The distribution of PS among the different regions of Brazil is very unequal, with the poorer North and Northeast regions having the lower PSWD (1.9 and 1.7 PS/100.000 children respectively). The worst distribution is seen on North Region where there is one state (Acre) that has only one registered pediatric surgeon for a population of approximately 900.000 inhabitants. This inequalities have been reported before and have a close relationship with the GDP/ capita^14^. Another study correlates the Brazilian unequal PSWD with mortality for children under 5 years^11^. Clearly, there is a need of governmental policies to address these disparities and to direct the allocation of surgical resources commensurate with local population needs.

Our findings, based on the expected incidences of pediatric surgical diseases, show that a Brazilian PS operates a total of 88 to 245 children/year (average 146 operations/year) depending on the region. However, a closer look on the surgical workload reveals that only 9 (6.1%) are high complexity (including neonatal) operations. Figure 3 reveals that the expected number of specific neonatal operations performed each year by the individual pediatric surgeon is very small, averaging less than one operation/year/pediatric surgeon, for instance, for esophageal atresia or diaphragmatic hernia.

These numbers are not exclusive for our country. The European Census on Pediatric Surgery showed that, in average, every EU surgeon performs 202 procedures/year, of which 11 procedures are on neonates^13^. A manpower study published in 2016 shows that in the US, many surgeons do not do any “complex index cases” for extensive periods. In that study, over the preceding year before re-certi□cation, the median number of cases per surgeon was one, for esophageal atresia repair, with almost 40% of surgeons doing none. Similarly, 40% of surgeons had not done any pull through operations for Hirschsprung’s Disease and 60% had not done any cases of either biliary atresia or choledochal cyst in the same period. ^15^. These data suggests that, keeping things the way they are, there maybe not enough cases for the average surgeon to maintain competence in the reconstructions necessary for the repair of complex congenital anomalies.

Although we admit that this is a very controversial issue, it would be desirable to start a national debate about be the best healthcare pathway to be offered to children in need of complex (especially neonatal) surgical care^2,16-18^. Care for these children is dependent not only on the presence of well trained and skilled pediatric surgeons but also on the existence of adequate facilities and highly specialized interdisciplinary pediatric teams (including pediatric anesthesiologists, neonatologists, radiologists, surgically trained nurses, etc.) to successfully face the challenges posed by these patients^19,20^

International experience suggests that the creation of decentralized regional centers in the different parts of the countries might not only improve the outcomes of these little patients but also rationalize the costs involved in their care^16,21-24^.

In 2008, APSA published a position statement saying that…”Because neonatal and infant surgical conditions are relatively uncommon and teams of appropriately skilled professionals and health systems properly resourced for expert perioperative care of infants are limited in number, the association strongly advocates that the surgical care of high-intensity infants occur within facilities with the human and institutional resources outlined. We view this approach as offering the greatest likelihood of providing optimal medical and surgical care to infants who have significant surgical conditions”^25^. Several studies confirmed that regionalization of neonatal surgical care is associated with improved surgical outcomes^26,27^.

In the UK, all biliary atresia patients are cared for in three centers: Leeds, Birmingham, and King’s College in London. Notably, there are a lot of well-known hospitals that are not on that list ^28^. The result is that each of the three centers in England sees more than 30 cases of biliary atresia per year. With that concentration, there has been an improvement in the transplant free survival, and the UK results now equal or surpass the best results in Europe^29,30^.

It is noteworthy that there is already an internal spontaneous “regionalization” within most pediatric surgery groups around the world, with some surgeons performing or supervising specific complex operations more often than others^31^. The development and growth of Pediatric Urology as a Pediatric Surgery subspecialty is probably one of the best examples of the spontaneous trend for subspecializacion, as is the evolution of Fetal Surgery^32^.

Arguments against regionalization of care are the eventual need for social dislocation for the family, the possibility of higher incidental costs and the possibility of pediatric surgeons losing their experience and potentially even income^16-18^.

However, experiences like, for instance, the Center for Anorectal Malformations or the Hepatobiliary Surgery in Cincinnati, among others, clearly show that concentrating the treatment of certain complex problems in resource-rich children’s environments may be better than approaching these problems as a once-in-a-year event in a general county hospital by a general pediatric surgeon. These centers have showed that, not surprisingly, most of the complicated cases they receive are children referred after failed attempts of primary surgical correction done in county hospitals by local pediatric surgeons who, theoretically, have been trained to treat those patients^33-35^.

Obviously, the main intention is not to restrict the practice of Pediatric Surgery in any center (which would be unethical), but rather to find ways to stimulate the development of resource-rich children’s environments devoted to the care of these children. In Brazil, the well-succeeded Liver Transplant Program administration model, where the eventual hospital that wishes to start a Liver Transplant Program has to comply to a fairly extensive list of standards in order to be accredited by the National Health Service and to receive payment for their services, might be a good starting point to discuss the creation of several regional resource-rich pediatric surgery centers, strategically distributed in the different regions of the country according to the expected number of patients and the existent workforce.

Just as important is the question on how do we plan the training of a Pediatric Surgeon to work in a situation where he or she will perform a fair number of low complexity operations a year but only one or two complex or neonatal surgeries? We are in a situation where the number of complex operations available for each training surgeon is decreasing, the time and cost of training a pediatric surgeon is increasing and the job market is changing drastically^36-39^. A survey among pediatric surgeons in the US shows that while the case load of “simple” procedures is increasing the number of complex operations is decreasing^40^. Apparently, most of the pediatric surgeons in Brazil today work in two or more different jobs, are constantly on call from two or more different hospitals and have little or no horizontal follow-up of the patients that they operate (personal communication). Although this is far from ideal care, this is the reality that the organization of health services in Brazil has lead us into. What are we doing to change this state of things? And, most importantly, how are we training the new pediatric surgeons to work in this scenario (or to be able to change it)?

If this is the scenario that will prevail in the near future, then maybe we should consider the possibility to train two levels of pediatric surgeons in Brazil. A general pediatric surgeon, that could be trained in two years (instead of the present three years) and a smaller number of specialized pediatric surgeons, selected among those who complete this general training, who would go through one or two more years of specialized training in the above-mentioned specialized centers, in order to be able to work with complex pediatric surgery in the high complexity pediatric surgery centers scattered around the country. It is believed that this could result in a rationalization of costs (training a Pediatric Surgery Fellow is a very expensive task) and also in better trained experts, improving the quality of the care offered to the children in need of complex surgical procedures^26^. Although controversial by nature, it is believed that this is a subject we should not refrain to examine carefully.

Data of the present study must be regarded with cautious because the results are based on the activities of the individual surgeon. It is known that in most pediatric hospitals, pediatric surgeons work together in variable size groups and so, the ideal scenario would be to be able to evaluate the workload of the pediatric surgery departments or services instead of the individual surgeons like it was done by the European Census for Pediatric Surgery^13^. This would result in a more realistic picture the present situation. Plans to perform such a study by means of a national survey are under way.

Also surgical expected workload might have been underestimated because it has been based on a list of index operations that does not include oncologic surgery, trauma, reoperations and the time to care for the children outside the operating room.

Another limitation of this study is the obvious disparity of information found, depending on the source one looks. In order to understand the problems and to plan the activities and the training programs of pediatric surgeons in Brazil it is paramount to know and understand the exact size and professional organization of the pediatric surgery workforce in the different regions of Brazil. For this to happen, it is imperative that standardization of the information is undertaken.

## Conclusion

Data presented in this paper shows that although the number of Pediatric Surgeons in Brazil appears to be adequate, there are profound regional differences that result in very unequal standards of care in the different regions of Brazil. Additionally, the expected workload of complex (including neonatal) operations is very small for each pediatric surgeon, making it necessary to discuss new policies to rationalize the care for children with complex surgical needs as well as the way we plan the training of new pediatric surgery residents.

This paper aims to offer a more grounded basis on which the future of Pediatric Surgery in Brazil should be planned. It is believed that this data may provide some help to all our colleagues, as well as national policy makers, to press for and to make better informed and well-grounded political choices in the field of pediatric surgery.

## Data Availability

All data produced in the present study are available upon reasonable request to the authors

https://datasus.saude.gov.br/informacoes-de-saude-tabnet/

